# Late Sleep Timing is Associated with Metabolic Risk in Postmenopausal Women

**DOI:** 10.64898/2026.10.02.26364600

**Authors:** J. Matthew Thomas, Philip A. Kern, Dorothy D. Sears, Samuel E. Armstrong, Cody Bumgardner, Catherine J. Russell, Courtney F. Murray, Jean L. Fry, Yuriko Katsumata, Julie S. Pendergast

**Affiliations:** Department of Biology, University of Kentucky, Lexington, KY, United States; Internal Medicine/Endocrinology, University of Kentucky, Lexington, KY, United States; College of Health Solutions, Arizona State University, Phoenix, AZ, United States; John Shufeldt School of Medicine and Medical Engineering, Arizona State University, Phoenix, AZ, United States; College of Medicine Office of Research, University of Kentucky, Lexington, KY, United States; Athletic Training and Clinical Nutrition, University of Kentucky, Lexington, KY, United States

**Keywords:** Circadian, visceral fat, accelerometry, obesity, insulin resistance

## Abstract

Postmenopausal women are vulnerable to metabolic dysfunction. Compelling evidence suggests this is because they lack the protective effects of estrogens. We have shown that circulating estrogens regulate daily eating and activity rhythms in female mice and protect them from obesity and metabolic dysfunction. However, few studies have investigated whether postmenopausal women have disrupted eating and sleep-activity rhythms that could contribute to metabolic dysfunction. The purpose of this cross-sectional study was to investigate the relationship between sleep and eating rhythms and metabolic risk in postmenopausal women with overweight or obesity. Seven days of sleep and food intake behavior data were collected from postmenopausal women with overweight or obesity. Sleep timing was assessed using wrist-worn actigraphy and sleep logs. First and last calorie intake times were collected daily using text messaging. Visceral adipose tissue (DXA), insulin resistance (Matsuda index), blood pressure, and triglycerides were analyzed as markers of metabolic risk. In 122 postmenopausal women (mean ± SD: 58.1 ± 3.6 yr, 33.2 ± 5.2 kg/m^2^ BMI), we found that later sleep onset was associated with greater visceral adipose tissue (*p*=0.0007), but not insulin resistance, blood pressure, or triglycerides. Next, we used a random-forest model to quantify the relative contributions of sleep and meal timing variables to predict visceral adipose tissue. The top predictors of visceral adipose tissue were time of sleep onset, sleep duration, and time of last caloric intake. These data suggest that interventions that advance sleep onset may reduce metabolic risk in postmenopausal women.

## Introduction

The circadian system coordinates 24-hr rhythms of physiology and behavior with environmental cycles^1^. Modern society has experienced a massive departure from reliance on environmental cycles to shape daily lives. Circadian misalignment, driven in part by sleeping and eating at inappropriate times of day, contributes to obesity and cardiometabolic risk^2-10^. Late-night eating and breakfast skipping are associated with obesity, insulin resistance, dyslipidemia, and cardiometabolic dysfunction.^2-10^. Likewise, later sleep timing is associated with higher BMI, body fat, abdominal obesity, and depression and anxiety^10-13^.

Sleep timing and meal timing are temporally interconnected behaviors, as individuals who sleep later also tend to eat later in the day^10^. While large epidemiological studies have linked sleep and meal timing to metabolic risk in a diverse population of adults, the relationship between sleep and eating rhythms and metabolic risk in postmenopausal women has been largely unexplored^2,4,6,12,14^. This population is important to study because metabolic risk increases dramatically following menopause.^15-17^. Compelling evidence suggests this is because they lack the protective effects of estrogens^18-20^. We showed that circulating estrogens regulate daily eating and activity rhythms in female mice and protect them from obesity and insulin resistance^19,20^. These findings suggest that the loss of circulating estrogens may impact circadian rhythms in women, underscoring the need to determine whether disrupted eating and sleep-activity rhythms contribute to metabolic dysfunction after menopause.

The menopausal transition is characterized by a rapid accumulation of visceral adipose tissue (VAT)^21,22^. VAT is strongly associated with dyslipidemia, hypertension, insulin resistance, cardiovascular disease, and type 2 diabetes and is a stronger predictor of cardiometabolic risk in women than in men^23-26^. Because VAT contributes to cardiometabolic disease after menopause, identifying modifiable behaviors associated with VAT is critical. To address this gap, we examined the association between sleep and meal timing and metabolic risk in postmenopausal women, with a focus on VAT and insulin resistance. We hypothesized that both late sleep and meal timing would be associated with increased VAT and insulin resistance.

## Materials and Methods

### Participants

Postmenopausal women between the ages of 45-65 years were recruited with social media and local advertisements as part of an ongoing clinical trial (NCT04893226). Participants completed in-person data collection at the University of Kentucky between October 2021 and September 2025. Cross-sectional baseline data from the clinical trial were analyzed for the current study once we reached recruitment of one-half of the planned enrollment for the baseline screening. Postmenopausal status was determined based on 1) participant self-report of ≥ 12 months since final menstrual period or 2) participant self-report of bilateral oophorectomy or 3) FSH ≥ 23 mIU/mL if ovaries were present and a procedure or medical condition permanently stopped menstruation (e.g., hysterectomy, uterine ablation). Since the goal of our study was to investigate sleep and meal timing in metabolically unhealthy postmenopausal women, we recruited women with obesity or overweight status with BMI ≥ 27 kg/m^2^. Exclusion criteria included participation in a weight loss program, change in body weight that exceeded 5% in the prior 3 months, participation in a new moderate to vigorous exercise regimen in the prior 3 months, uncontrolled sleep disorders, taking melatonin or other medications or supplements for the purpose of promoting or inhibiting sleep, significant psychiatric disorders, diagnosed diabetes, diagnosed coronary artery disease, diagnosed dysregulated eating behaviors, having care-taking responsibilities that significantly impact sleep, regularly consuming more than 2 alcoholic beverages per day, taking medications for weight loss or medications that impact appetite, smoking more than 5 cigarettes/day or having a greater than or equal to 30 pack-year history of smoking, working night shift or rotating shift work within the past 6 months, taking systemic hormone therapy (estradiol/progesterone), previous abdominal surgeries for weight loss, taking selective estrogen-receptor modulators, practicing intermittent fasting or time-restricted eating, and logging food intake for the purposes of weight loss. Two participants were excluded from analysis because of abdominal surgeries for weight loss. One participant was excluded from analysis because of taking a dual GIP and GLP-1 receptor agonist and one participant was excluded from analysis because of taking a selective estrogen-receptor modulator. One participant withdrew from the study before completion and was excluded from analysis.

Participants were informed of the purpose of the study and provided written informed consent prior to participation. Medical history and medications were recorded for each participant.

Participants completed the International Physical Activity Questionnaire (IPAQ) to assess physical activity level and the STOP-Bang questionnaire to assess risk for obstructive sleep apnea^27^. Study method and results are reported following the Strengthening the Reporting of Observational Studies in Epidemiology (STROBE) Statement for cross-sectional studies^28^.

### Sleep

Participants wore a triaxial actigraphy device (Actigraph wGT3X-BT, Ametris, Pensacola, FL) on the nondominant wrist and completed sleep logs for 7 days (**Figure 1**). One participant was excluded from analysis due to missing actigraphy and sleep log data. One participant wore the actigraphy monitor on dominant wrist due to a wrist brace on the non-dominant wrist. Actigraphy data was downloaded and reintegrated into 60 second epochs and analyzed using the ActiLife software and the Cole-Kripke algorithm^29^. Sleep logs were used to help identify in-bed and out-of-bed times for participants. Sleep outcomes included average sleep onset, wake time, sleep duration, and sleep efficiency. Sleep efficiency was calculated as: (Total time scored as sleep/Total time in bed trying to sleep) x 100.

**Figure 1.**
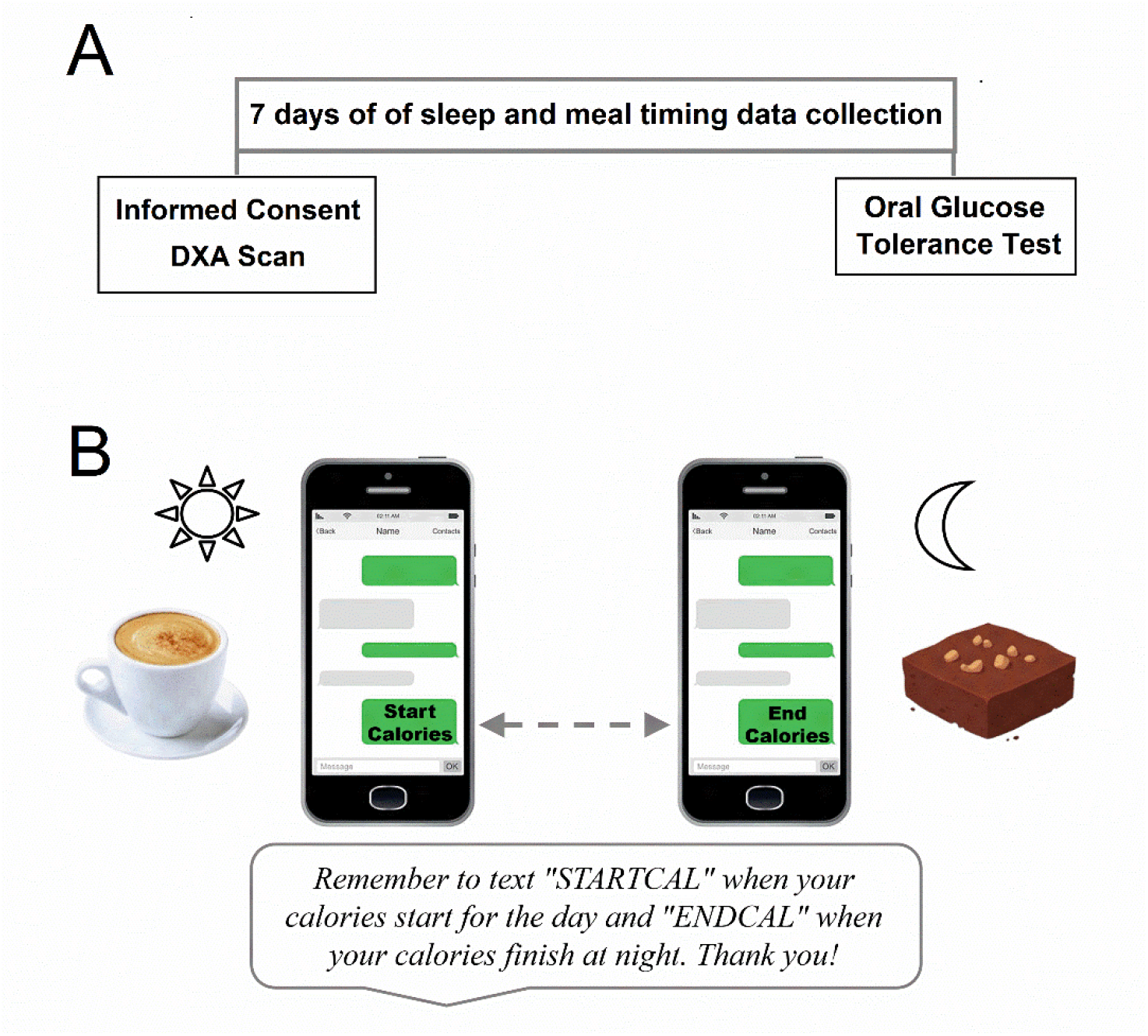
Study Protocol and text messaging system. During consent, anthropometric measures were collected, participants completed a DXA scan, and participants began wearing an actigraphy monitor and recording in sleep logs (**A**). Participants recorded daily meal timing for 7 days using an automated text-messaging system (**B**).

### Meal timing

Participants recorded first and last calorie intake times each day with a state-based text messaging system that we developed (**Figure 1**)^30^. Daily scheduled reminders prompted participants to send data each day if no message was received by the study team. Daily calorie window was calculated as the duration between first caloric intake and last caloric intake.

Consistent with a previous study, caloric events recorded before 4 a.m., and in the middle of a sleep episode, were classified as within the prior day’s eating window^10^. Time of caloric intake relative to sleep onset and wake time was also calculated.

### Body composition and blood pressure

Height was measured to the nearest 0.1cm and body mass to the nearest 0.1kg and used to calculate BMI (kg/m^2^). Five participants were excluded from analysis due to a low BMI (<27 kg/m^2^). Waist circumference was collected in triplicate at the level of the umbilicus and the average value was used for analysis. Body composition was measured using total body DXA scans (Lunar iDXA, GE Healthcare, Chicago, IL). Body fat percentage was collected for each participant, and visceral adipose tissue mass was estimated with the Enhanced CoreScan Feature. Blood pressure was collected in triplicate (Welch Allyn Connex Vital Signs Monitor, Hillrom, Deerfield, IL) and the average value was used for analysis.

### Fasting labs

Participants provided a fasting blood sample, and the following laboratory panels were analyzed by Quest Diagnostics: complete metabolic panel, complete blood count, hemoglobin A1c (HbA1c), lipid panel, thyroid stimulating hormone (TSH), and follicle stimulating hormone (FSH; if necessary to confirm postmenopausal status). Three participants were excluded from analysis based on a premenopausal level of FSH measured from fasting labs. Fasting triglycerides were included in the analyses for this study.

### Insulin Sensitivity

Each participant completed an oral glucose tolerance test (OGTT). Briefly, participants consumed a 75g glucose beverage (75 SUN-DEX, Fisherbrand, Waltham, MA). Blood samples were collected every 30 minutes following the glucose beverage with the last sample collected at 2 hours. Plasma glucose was measured with a YSI biochemistry analyzer (2900D, Xylem, Yellow Springs, Ohio) and plasma insulin was measured using a U-PLEX Human Insulin Assay (K1516HK-2, Meso Scale Diagnostics, Rockville, MD). Glucose and insulin concentrations from all OGTT timepoints were used to calculate Matsuda index as an estimate of total body insulin sensitivity^31^. Two participants met 3 criteria for diabetes from the OGTT and were excluded from analysis. Matsuda index was not calculated for two participants because not all samples were collected during the OGTT because of technical difficulties during blood draws, and for one participant because of excess hemolysis affecting insulin measurements.

### Statistical Analyses

Data were analyzed using R, version 4.5.3. Descriptive data are presented as means ± standard deviation. Welch t-tests were used to assess differences in sleep timing, sleep duration, first and last caloric intake, and calorie window between participants with overweight (BMI < 30) and obese (BMI ≥ 30) status. Pearson’s method was used to estimate the correlations between sleep and eating rhythm variables and visceral adipose tissue and Matsuda index. Furthermore, since the accumulation of visceral adipose tissue is strongly associated with dyslipidemia and hypertension, we also included fasting triglycerides and systolic and diastolic blood pressure in our Pearson correlation analysis^32-35^. We used a Bonferroni-corrected significance level of 0.00125 for the Pearson correlation analyses to minimize type 1 error. Next, we used a random-forest model to quantify the relative contributions of sleep and meal timing variables to predict visceral adipose tissue mass (randomForest R package^36^, version 4.7.1.2). The random forest model was constructed using 10,000 decision trees and the variable importance was assessed using increase in node purity. We next conducted a multivariable linear regression analysis to examine the association between sleep onset time and visceral adipose tissue, adjusting for sleep duration and timing of the last calorie intake.

## Results

One hundred thirty-eight participants were enrolled for the study. After exclusion of 16 participants (Supplemental Figure S1), 122 were included in the analytic dataset (**Table 1**). Forty-four participants (36.1%) had two criteria of the metabolic syndrome, and 49 (40.2%) participants had the metabolic syndrome, defined as having ≥3 metabolic syndrome criteria. Ninety participants (73.8%) were classified as having prediabetes based on American Diabetes Association (ADA) criteria (HbA1c 5.7–6.4%, fasting glucose 100–125 mg/dL, or 2-hour OGTT 140–199 mg/dL)^37^. Although no participants in our analytic dataset had been diagnosed as diabetic, 9 participants (7.4%) met 1 criterion for diabetes and 2 participants (1.6%) met 2 criteria for diabetes, based on ADA criteria (fasting plasma glucose ≥126 mg/dL, HbA1c ≥6.5%, 2-hour plasma glucose ≥200 mg/dL). Twenty-nine participants were taking an antihypertensive medication.

**Table 1.** Characteristics of study participants (N=122)*

| Variable | Mean ± SD<br>(range) |
| --- | --- |
| Age (yr) | 58.1 ± 3.6<br>(49.0-65.0) |
| **Anthropometric and body composition** |  |
| BMI (kg/m<sup>2</sup>) | 33.2 ± 5.2<br>(26.6-50.6) |
| Waist circumference (cm) | 108.9 ± 12.6<br>(88.0-152.1) |
| Body fat (%) | 46.1 ± 5.3<br>(35.3-62.2) |
| Visceral adipose tissue (kg) | 1.5 ± 0.7<br>(0.4-4.3) |
| **Metabolic risk** |  |
| HbA1c (%) | 5.6 ± 0.3<br>(5.0-6.5) |
| HDL (mg/dL) | 60.9 ± 13.7<br>(34.0-103.0) |
| Triglycerides (mg/dL) | 117.7 ± 50.4<br>(48.0-319.0) |
| Matsuda index | 2.4 ± 1.7<br>(0.3-9.0) |
| **Sleep** |  |
| Sleep onset | 23:07 ± 01:01<br>(20:26-02:13) |
| Sleep efficiency (%) | 89.2 ± 4.1<br>(73.5-97.7) |
| Wake time | 6:39 ± 01:00<br>(4:32-9:34) |
| Sleep duration (h) | 7.5 ± 0.9<br>(4.8-10.1) |
| **Meal timing** |  |
| Time of first caloric intake | 8:15 ± 01:22<br>(5:15-11:37) |
| Time of last caloric intake | 20:27 ± 01:13<br>(18:00-23:41) |
| Daily calorie window (h) | 12.2 ± 1.6<br>(7.3-15.7) |
| Time from wake to first calorie (h) | 1.6 ± 1.2<br>(0.1-6.1) |
| Time from last calorie to sleep (h) | 2.7 ± 1.1<br>(-0.3 – 6.6) |
| *One hundred thirty-eight participants were enrolled for the study. After exclusion of 16 participants (Supplemental Figure S1), 122 were included in the analytic dataset. |  |

Average sleep onset, wake time, sleep duration, and sleep efficiency were assessed over 7 days using actigraphy and sleep logs (**Figure 2**). Average time of sleep onset was significantly later in participants with obesity (N=83) compared to participants with overweight status (N=39) (*p*=0.01; **Figure 2**). Average wake time and sleep duration did not differ between participants with obesity and participants with overweight status. Average sleep efficiency for the cohort was 89.2%, which is above the clinical threshold for good sleep quality (≥85%)^38^.

**Figure 2.**
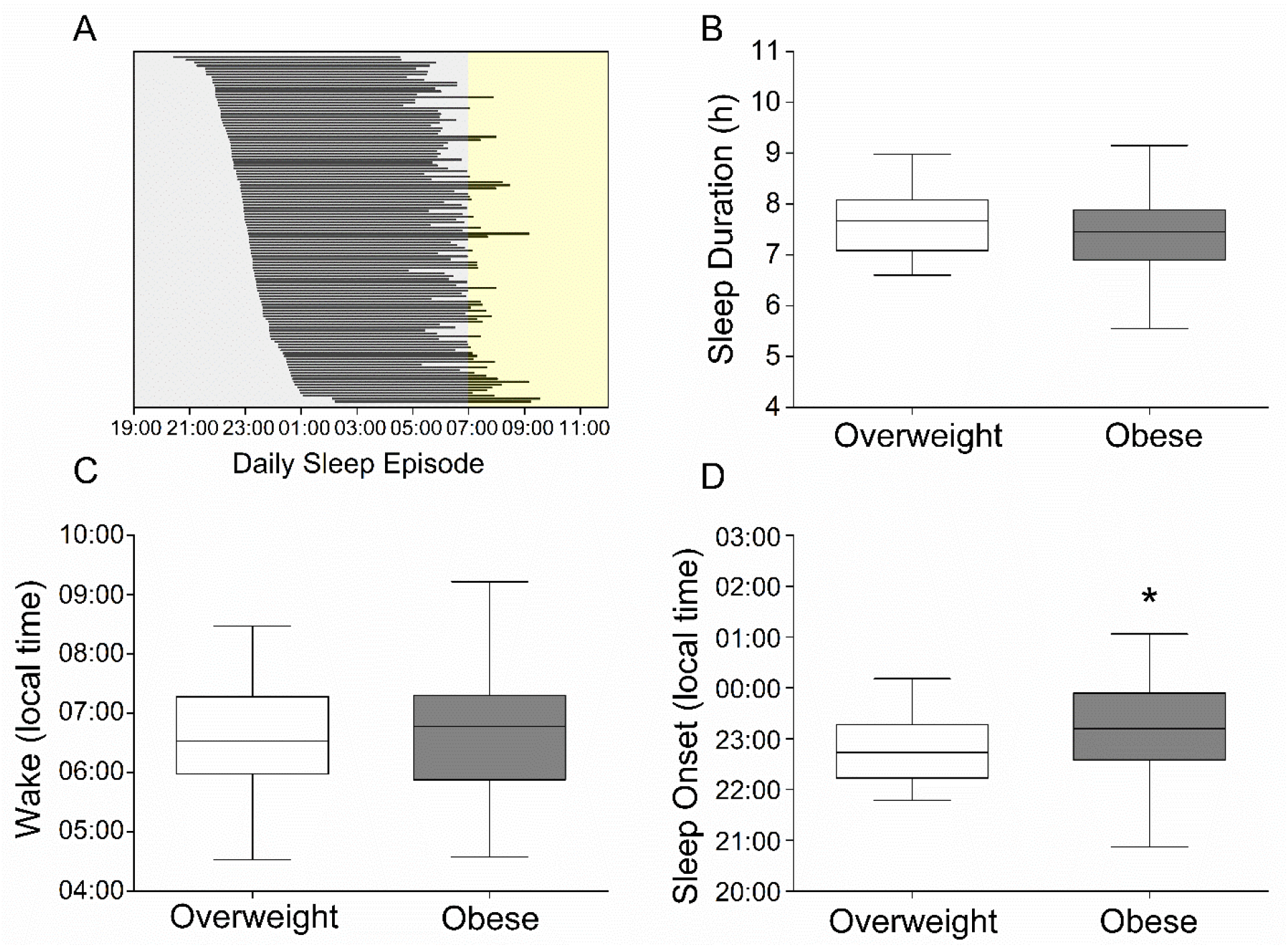
Sleep timing in postmenopausal women. (**A**) Horizonal bars show the average sleeping window for each participant (n=122). Bars are sorted by sleep onset time. (**B**) Sleep duration, (**C**) wake time, and (**D**) sleep onset in participants with obesity or overweight status were analyzed using Welch’s t-test. \**p*<0.05

Average times of first and last caloric intake for 7 days were used to assess meal timing and daily calorie window (**Figure 3**). Average daily calorie window in our sample was 12.2 h, and 76 participants (62%) consumed their last caloric intake after 8:00pm. We found no differences in average timing of first caloric intake, last caloric intake, or daily calorie window between participants with obesity versus participants with overweight status.

**Figure 3.**
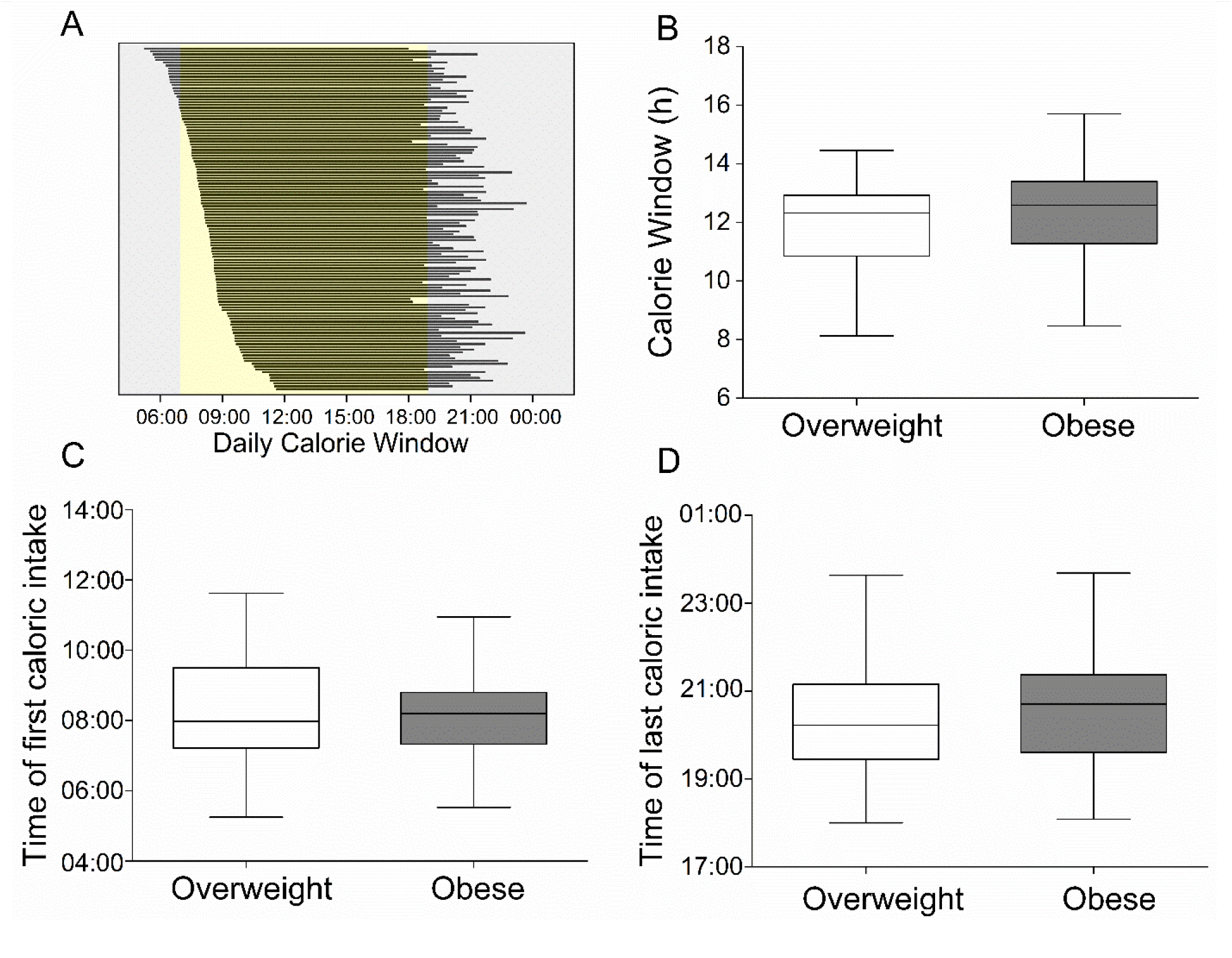
Meal timing in postmenopausal women. (**A**) Horizonal bars show the average eating window for each participant (n=122). Bars are sorted by time of first caloric intake. (**B**) Calorie window, (**C**) time of first caloric intake, and (**D**) time of last caloric intake in participants with obesity or overweight status were analyzed using Welch’s t-test (all *p*>0.05).

Next, we analyzed the association between sleep and meal timing variables with metabolic risk outcomes (**Table 2**). We found that later sleep onset was associated with greater visceral adipose tissue (r=0.30; *p*=0.0007; Bonferroni corrected α=0.00125). None of the other sleep or meal timing variables was associated with visceral adipose tissue, blood pressure, triglycerides, or Matsuda index (**Table 2**). In exploratory analysis, we found that later last caloric intake was associated with greater waist circumference (**Supplemental Table S1**), which is a surrogate anthropometric measure of visceral adipose tissue^39^.

**Table 2.** Pearson correlations coefficients among sleep and eating rhythms and metabolic risk.

|  | Visceral<br>adipose<br>tissue mass | Matsuda<br>index | Systolic<br>blood<br>pressure | Diastolic<br>blood<br>pressure | Triglycerides |
| --- | --- | --- | --- | --- | --- |
| Sleep onset | 0.30* | -0.04 | -0.14 | -0.07 | 0.06 |
| Wake | 0.17 | -0.02 | -0.02 | 0.11 | 0.07 |
| Sleep duration | -0.16 | 0.03 | 0.13 | 0.20 | 0.01 |
| Time of first calorie | 0.09 | 0.00 | -0.08 | 0.03 | -0.04 |
| Time of last calorie | 0.20 | 0.02 | -0.13 | -0.02 | -0.04 |
| Daily caloric window | 0.08 | 0.02 | -0.04 | -0.04 | 0.01 |
| Wake to first calorie | -0.04 | 0.02 | -0.08 | -0.07 | -0.12 |
| Last calorie to sleep | 0.06 | -0.06 | 0.02 | -0.04 | 0.10 |

\* $p < 0.00125$

Next, we determined which sleep and meal timing variables were strongest predictors of visceral adipose tissue mass (**Figure 4**). Sleep onset, followed by sleep duration, and time of last caloric intake were the strongest predictors of visceral adipose tissue mass. In contrast, daily caloric window was the weakest predictor of visceral adipose tissue. Lastly, we found that a one-hour delay in sleep onset was associated with a 0.18 kg increase in visceral adipose tissue after adjusting for sleep duration and timing of last calorie intake (*p*=0.02).

**Figure 4.**
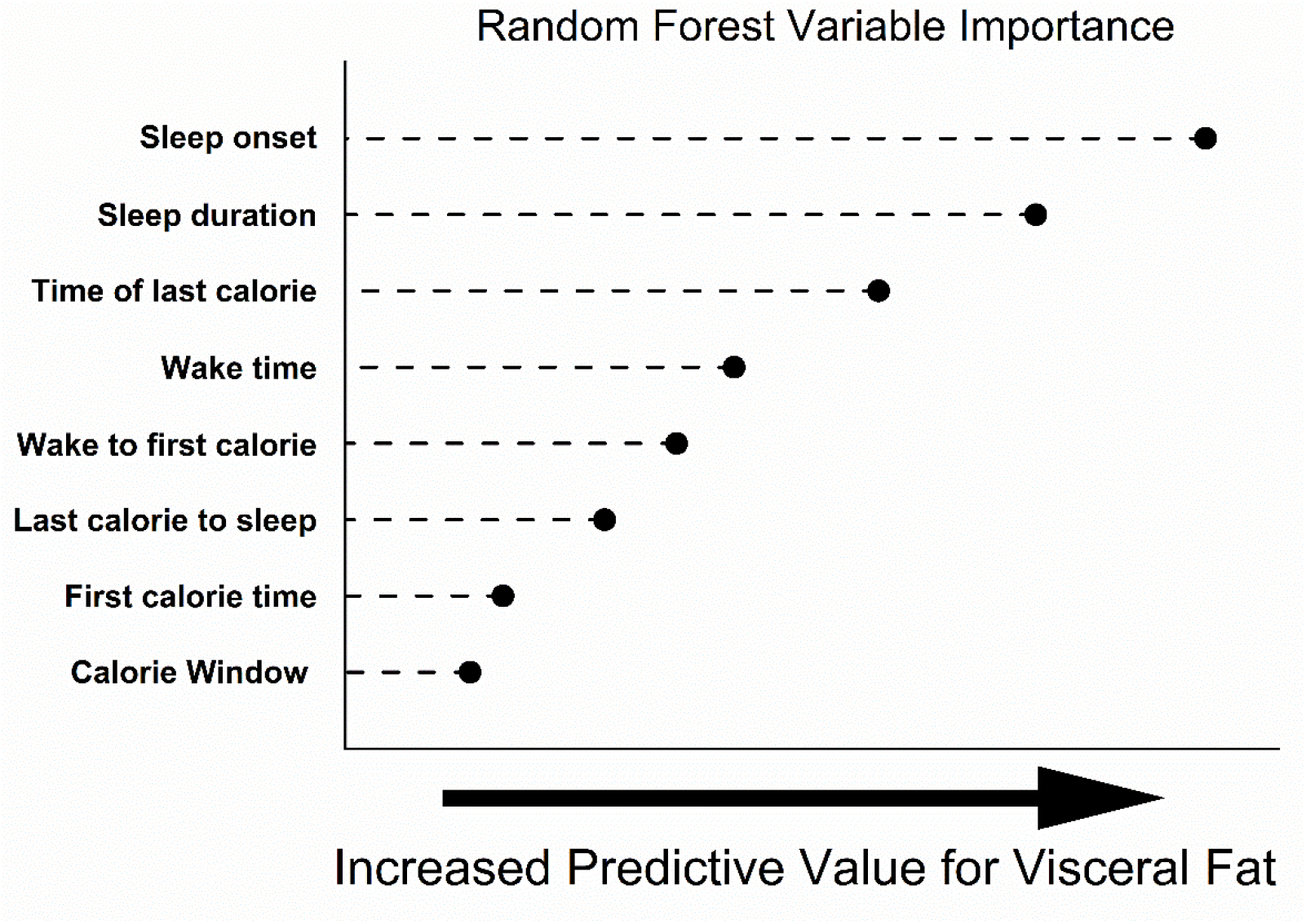
Sleep onset predicts visceral adipose tissue in postmenopausal women. Predicitve value for sleep and meal timing variables plotted using node purity values. Data were analyzed using a random forest model constructed using 10,000 decision trees.

## Discussion

In this study, we examined the relationships between sleep and meal timing and markers of metabolic risk in postmenopausal women. We found that later sleep timing was a predictor of visceral adipose tissue in postmenopausal women. These findings are consistent with large cross-sectional studies of middle age and older adults reporting that later sleep timing is associated with a greater risk of obesity^12,40^. Our findings are novel because relatively few studies have examined these behaviors specifically in postmenopausal women.

The limited research on sleep characteristics and metabolic risk in postmenopausal women has largely focused on sleep duration and sleep quality and obesity. For example, short sleep duration and poor sleep quality are associated with dyslipidemia, insulin resistance, and larger waist circumference in this population^41-44^. However, the timing of sleep, which may be a more promising target for behavioral interventions, has remained understudied in postmenopausal women.

The relationship between later sleep timing and greater visceral adipose tissue may be explained, in part, by chronic circadian misalignment. The circadian system coordinates 24-hour rhythms of physiology and behavior and entrains or synchronizes these rhythms to the light/dark cycle^1^. A late sleep onset creates misalignment between the circadian system, the light/dark cycle, and sleep/wake behavior. Laboratory and epidemiological studies show that circadian misalignment contributes to glucose intolerance, reduced insulin sensitivity, increased blood pressure, increased inflammatory markers, increased obesity, fat mass, and risk of the metabolic syndrome^45-48^. Studies show that late sleep timing is also associated with less healthy behaviors, such as poor dietary choices and sedentary behavior, that contribute to obesity^49-51^. Consistent with other studies in middle age and older adults, we did not find an association between wake time and metabolic risk^12,52^. In contrast to previous studies, we found no association between sleep duration and metabolic risk. Although limited, other studies in postmenopausal women show that short sleep duration is associated with larger waist circumference and a greater diabetes risk^44,53^. In our dataset only four women had sleep duration less than six hours, which is a common threshold for increased metabolic risk^44,54,55^. Therefore, our narrow distribution of sleep duration may have influenced our statistical power for this analysis.

In our study of postmenopausal women, sleep efficiency, which is a measure of sleep quality, was above the clinical threshold for good sleep quality (>85%)^38^. Although many studies report poor subjective sleep quality among postmenopausal women, evidence from polysomnography, the gold standard for assessing sleep quality, is inconclusive when comparing premenopausal and postmenopausal women^56-59^. Average sleep efficiency from polysomnography in a study of obese postmenopausal women was 76%, indicating poor sleep quality^60^. Therefore, some awakenings may not have been detected with actigraphy, and therefore the sleep efficiency from our cohort may be overestimated.

We also found that the times of first and last caloric intake, as well as daily calorie window, were not associated with visceral adipose tissue or insulin resistance in this cohort. However, in an exploratory analysis, we found that later time of last caloric intake was associated with greater waist circumference, which is a surrogate anthropometric measure of visceral adipose tissue^39^. In the current study we investigated the time of caloric intake relative to both clock time and the sleep/wake cycle as a proxy of circadian phase. Studies using both approaches in other populations consistently show that late night food intake is associated with increased obesity and early morning food intake is associated with lower odds of obesity^5,6,8^. Furthermore, time-restricted eating studies show that restricting food intake to earlier in the day improves insulin resistance, blood pressure, and body weight and visceral adipose tissue in participants with obesity, metabolic syndrome, or prediabetes^61-63^.

A strength of our study is the characterization of a cohort of postmenopausal women with assessment of metabolic risk using body composition and a robust measure of insulin resistance. While previous studies examined the relationship between sleep and meal timing and obesity, measured by BMI, our study estimated visceral adipose tissue with a DXA scan, which is a more direct measure of metabolic risk^5,39^. Visceral adipose tissue also dramatically increases following the menopausal transition and is a stronger predictor of cardiometabolic risk in women compared to men^21,23,24,64^. Each 1-kg increase in visceral adipose tissue is associated with a 7-fold increased odds of type 2 diabetes in women compared to just a 2-fold increase in men^24^. A previous study found that a threshold of 1.1kg of visceral adipose tissue is an indicator of elevated metabolic risk in older women^65^. Seventy percent of the women in this study were above this threshold, indicating that this was a metabolically unhealthy cohort. We found that every 1-hour increase in sleep onset was associated with an increase in visceral adipose tissue. Our results suggest that a late sleep timing is a behavior that may be targeted to decrease metabolic risk^66^.

There were some limitations of our study. 1) Our analysis of the timing of the first and last caloric intake during the study did not account for the caloric content or diet composition of the caloric intake. Studies show that the proportion of daily caloric intake and the macronutrient content of nighttime food intake may influence the effect of meal timing on obesity, however the influence of specific macronutrients is mixed^5,67^. 2) The gold standard methods for measurement of visceral adipose tissue are X-ray computed tomography (CT) and magnetic resonance imaging (MRI). However, we used a DXA scan to estimate visceral adipose tissue, which is highly correlated with CT and MRI scan measurements and safe for participants due to the low radiation dose^68^.

In summary, we found that later sleep timing predicts increased metabolic risk in postmenopausal women with overweight and obesity. This population is important to study because metabolic risk increases dramatically following menopause.^15-17^ These data suggest that interventions that advance sleep onset may be a promising target to improve metabolic risk in postmenopausal women. For example, increased morning light exposure advances circadian phase and sleep onset^69,70^. Recent studies show that time-restricted eating, which has been studied primarily as an intervention to improve obesity and metabolic risk, can also advance sleep and improve sleep quality^71-74^. Future studies should investigate whether interventions that advance sleep timing can improve metabolic risk in postmenopausal women.

## Data Availability

All data produced in the present study are available upon reasonable request to the authors.

## Data Availability Statement

The data underlying this article will be shared on reasonable request to the corresponding authors.

