## Supplementary material for "Late Sleep Timing is Associated with Metabolic Risk in Postmenopausal Women": Table S1

| **Supplemental Table S1. Pearson correlations coefficients among sleep and eating rhythms and obesity and metabolic risk outcomes** | | | | | |
| --- | --- | --- | --- | --- | --- |
|  | BMI | Waist circumference | Body fat percentage | HDL | Fasting glucose |
| Sleep Onset | r=0.178 | r=0.284 | r=0.317* | r=-0.105 | r=0.209 |
|  | *p*=0.050 | *p*=0.002 | *p*<0.000 | *p*=0.252 | *p*=0.021 |
| Wake | r=0.056 | r=0.105 | r=0.250 | r=0.054 | r=0.096 |
|  | *p*=0.537 | *p*=0.250 | *p*=0.005 | *p*=0.551 | *p*=0.294 |
| Sleep duration | r=-0.143 | r=-0.212 | r=-0.083 | r=0.187 | r=-0.135 |
|  | *p*=0.116 | *p*=0.019 | *p*=0.362 | *p*=0.040 | *p*=0.139 |
| Time of first calorie | r=0.053 | r=0.013 | r=0.170 | r=0.115 | r=0.054 |
|  | *p*=0.565 | *p*=0.890 | *p*=0.062 | *p*=0.209 | *p*=0.551 |
| Time of last calorie | r=0.249 | r=0.299* | r=0.242 | r=0.026 | r=0.048 |
|  | *p*=0.006 | *p*<0.001 | *p*=0.007 | *p*=0.779 | *p*=0.600 |
| Daily caloric window | r=0.148 | r=.221 | r=0.041 | r=-0.079 | r=-0.015 |
|  | *p*=0.104 | *p*=0.014 | *p*=0.656 | *p*=0.389 | *p*=0.870 |
| Wake to first calorie | r=0.014 | r=-0.076 | r=-0.015 | r=0.088 | r=-0.018 |
|  | *p*=0.882 | *p*=0.408 | *p*=0.865 | *p*=0.334 | *p*=0.842 |
| Last calorie to sleep | r=-0.113 | r=-0.068 | r=0.025 | r=-0.125 | r=0.140 |
|  | *p*=0.216 | *p*=0.454 | *p*=0.787 | *p*=0.170 | *p*=0.123 |
| **p*<0.00125 |  |  |  |  |  |
