## Supplementary figures and images for "Late Sleep Timing is Associated with Metabolic Risk in Postmenopausal Women"

### Figure S1

**Supplemental Figure S1. STROBE flow diagram for enrollment of study participants**

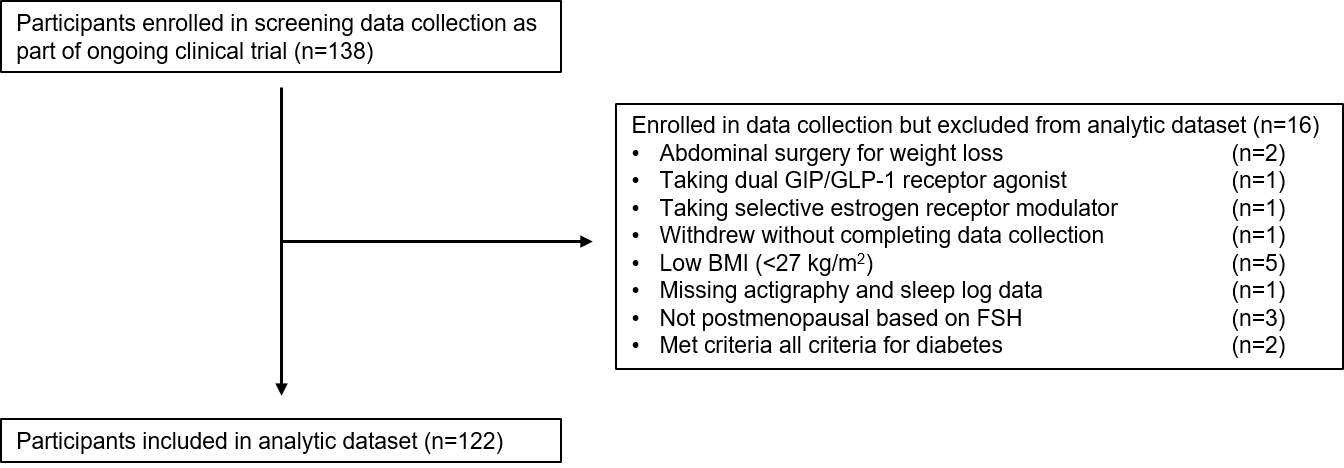
